# Acute myelitis after SARS-CoV-2 infection: a case report

**DOI:** 10.1101/2020.03.16.20035105

**Authors:** Kang Zhao, Jucun Huang, Dan Dai, Yuwei Feng, Liming Liu, Shuke Nie

**Author notes:** Corresponding authors: Liming Liu, Address: Zhongshan Road 26, Renmin Hospital of Wuhan University, Wuhan 430033, China, Shuke Nie, Address: Jiefang Road 238, Renmin Hospital of Wuhan University, Wuhan 430060, China. **Disclosure:** The authors declare no financial relationships with any organizations that might have an interest in the submitted work. The authors declare no competing interests.

## Abstract

We report a case of acute myelitis in a patient infected with severe acute respiratory syndrome coronavirus 2 (SARS-CoV-2). A 66-year-old man with coronavirus disease 2019 was admitted with acute flaccid paralysis of the bilateral lower limbs and urinary and bowel incontinence. All serum microbiological test results were negative, except for SARS-CoV-2 nucleic acid testing. Clinical findings indicated post-infectious acute myelitis. He received treatment containing ganciclovir, lopinavir/ritonavir, moxifloxacin, dexamethasone, human immunoglobulin, and mecobalamin. With a diagnosis of post-infectious acute myelitis and comprehensive treatment, paralysis of the bilateral lower extremities ameliorated. After two negative novel coronavirus RNA nasopharyngeal swab tests, he was discharged and transferred to a designated hospital for isolation and rehabilitation therapy.

## Background

Coronavirus disease 2019 (COVID-19) was first reported in Wuhan, China in December 2019^[1]^. It has been confirmed that this disease is caused by a new type of enveloped RNA coronavirus B, which is named severe acute respiratory syndrome (SARS) coronavirus 2 (SARS-CoV-2)^[2]^. As of April 3, 2020, there were 1017914 laboratory-confirmed cases worldwide, resulting in 53188 deaths. Studies have shown that the virus has 79% homology with the SARS virus^[3]^, and the potential intermediate animal host remains unknown, although it has been demonstrated that this virus did not originate directly from pangolins^[4]^. Researchers have confirmed that SARS-CoV-2 enters the human body through angiotensin-converting enzyme 2 (ACE2) receptors on the surface of human cells and causes disease^[5]^. ACE2 is expected to be a possible target for intervention and treatment of the disease. ACE2 receptors are found in type II alveolar epithelial cells of the human lungs; therefore, it has become the main target of SARS-CoV-2 in the pathogenesis of COVID-19^[6]^. However, in the processes of clinical diagnosis and treatment, it has been found that many critically ill patients exhibit symptoms of multiple organ dysfunction, in addition to lung, liver, and kidney damage being very prominent, which may be related to the expression of ACE2 in hepatic bile duct cells and proximal renal tubules^[7,8]^.

ACE2 receptors are also expressed on the surface of spinal cord cells^[9,10]^. Whether spinal cord neurons are implicated in COVID-19 remains unknown. We report a patient with COVID-19 who suddenly developed acute myelitis after an initial high fever, suggesting that the central nervous system (CNS) may also be attacked by SARS-CoV-2.

### Case presentation

A 66-year-old man was admitted to the hospital with fever and fatigue for 2 days in Wuhan, China. He had no contact with patients with COVID-19. He developed a fever without an obvious cause on February 8, 2020, and his highest body temperature was 39°C, with fatigue and without cough, asthma, and dyspnea. He visited the outpatient clinic of a local hospital in Wuhan and was administered oral moxifloxacin hydrochloride and oseltamivir for 5 days. On February 13, 2020, he underwent chest computed tomography (CT) in the local lung hospital, which revealed patchy changes in both lungs (Figure 1A, B). The novel coronavirus (nCoV) RNA nasopharyngeal swab test result was positive; he was diagnosed with mild COVID-19. He was admitted to Wuhan Cabin Hospital and treated in individual isolation. After a high fever (40°C) at night, he developed weakness in both lower limbs with urinary and bowel incontinence, culminating in flaccid lower-extremity paralysis. His condition deteriorated rapidly, and he was transferred to the intensive care unit for critical care and treatment. Examination of his vital signs revealed the following results: oxygen saturation, <93% at rest; body temperature, 37°C; pulse, 80 bpm; respiratory rate, 18 breaths/min; and blood pressure, 81/51 mmHg. No obvious abnormality was found on cranial nerve examination. His neurologic examination demonstrated 3/5 power with normal reflexes in the bilateral upper extremities and 0/5 power reduction with hyporeflexia in the bilateral lower extremities. Sensation was intact in all modalities in the arms but was globally impaired in both legs. There was a sensory level at T10 to pinprick testing, with feelings of paresthesia and numbness below the level. The tendon reflexes of both lower limbs decreased, and pain, temperature, and tactile sensations decreased below the level of chest 10. Bilateral pathological signs were negative.

**Figure 1.**
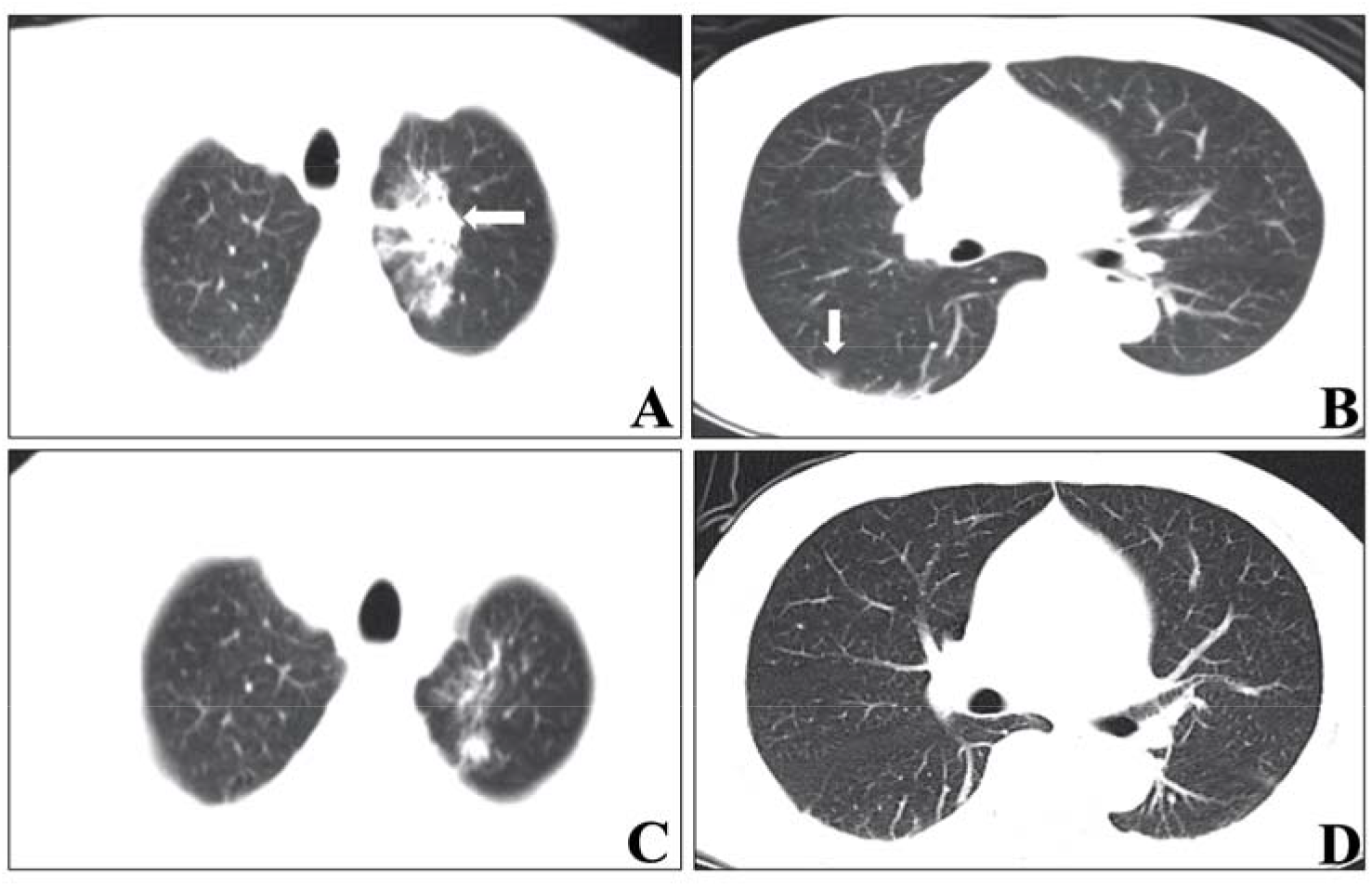
Chest CT images of the patient with COVID-19. Chest CT showed a patchy, high-density, blurred shadow in the upper lobe of the left lung (A) and patchy ground-glass shadow in the anterior segment of the upper lobe of the right lung (B). After treatment, chest CT showed that the previous lesions were almost completely absorbed. CT, computed tomography.

The routine blood, C-reactive protein, serum amyloid protein, procalcitonin, and large biochemistry test results after admission are shown in Table 1. On admission, the carcinoembryonic antigen level was normal, serum ferritin level was >2000 (normal range 21.81–274.6 ng/mL), and electrolytes showed no obvious abnormality, except serum iron (Table 1). Hypersensitive troponin levels were normal. Blood lipids showed low high-density lipoproteinemia, a high-density lipoprotein cholesterol (HDL-C) level of 0.51 (1.16–1.42 mmol/L), and interleukin-6 level of 56.72 (0–7 pg/mL). Among pathogenic microbes, the immunoglobulin M (IgM) of *Chlamydia pneumoniae*, Epstein–Barr virus (EBV) antibody, influenza B virus, adenovirus, coxsackievirus, *Mycoplasma pneumoniae*, influenza A virus, parainfluenza virus, cytomegalovirus (CMV), and respiratory syncytial virus all tested negative. T cells of tuberculosis infection were negative. The results of the nCoV RNA nasopharyngeal swab are shown in Table 2. Before treatment on February 15, 2020, chest CT showed a patchy high-density blurred shadow in the upper lobe of the left lung (Figure 1A) and patchy ground-glass shadow in the anterior segment of the upper lobe of the right lung (Figure 1B), suggesting viral pneumonia. Cranial CT revealed bilateral basal ganglia and paraventricular lacunar infarction and brain atrophy (Figure 2). Cerebrospinal fluid serological testing and magnetic resonance imaging of the spinal cord were not performed for pandemic-related reasons during hospitalization.

**Table 1.**
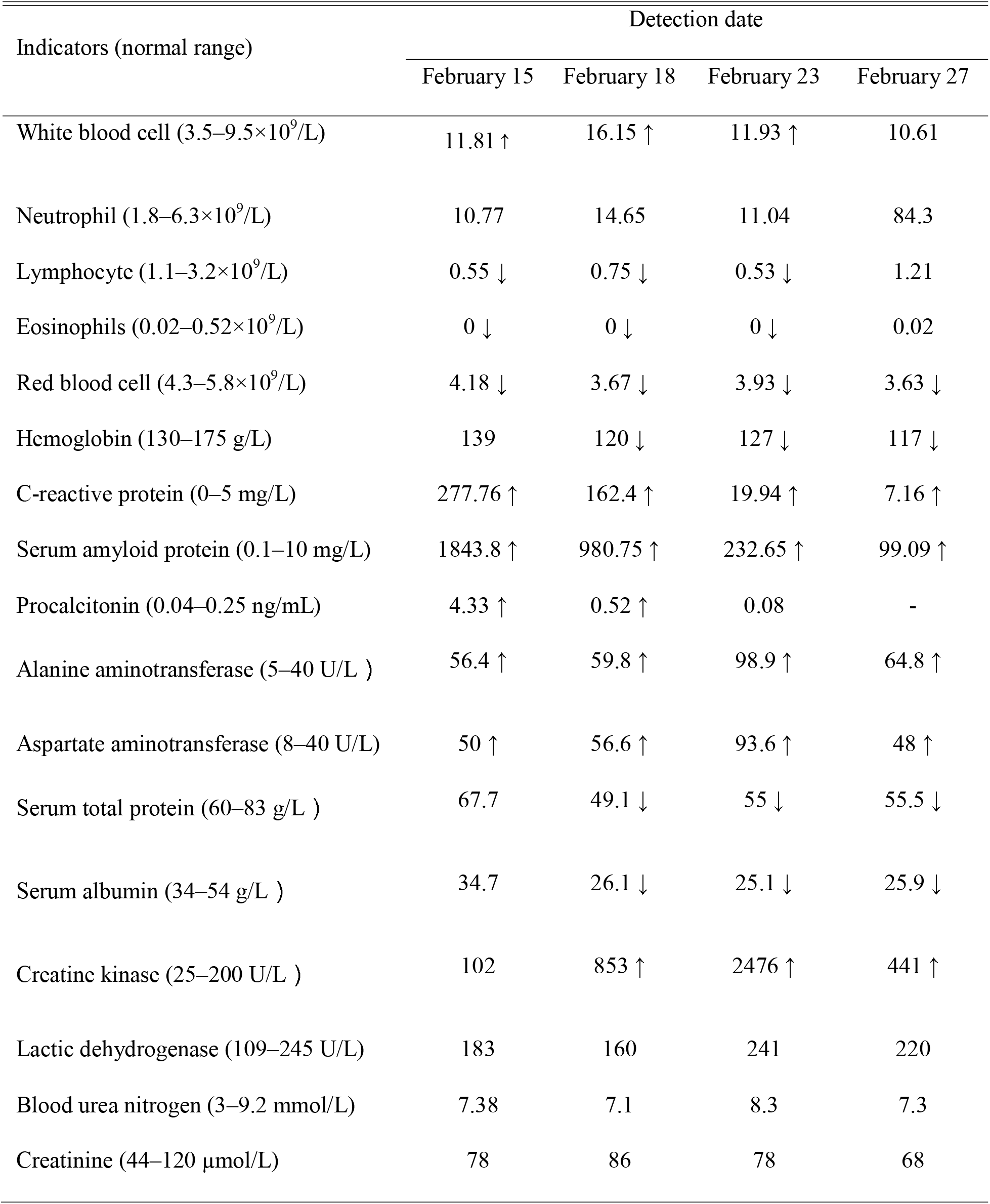
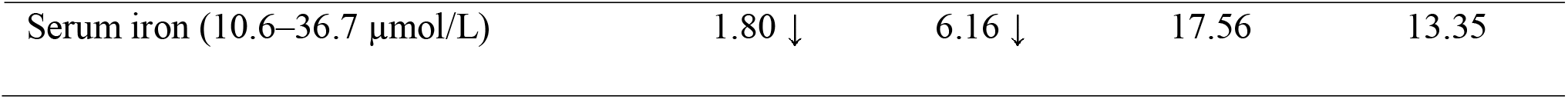
**Results of blood routine and biochemical tests after admission**

| Indicators (normal range) | Detection date |  |  |  |
| --- | --- | --- | --- | --- |
|  | February 15 | February 18 | February 23 | February 27 |
| White blood cell ($3.5\text{--}9.5\times 10^9/\text{L}$) | 11.81 ↑ | 16.15 ↑ | 11.93 ↑ | 10.61 |
| Neutrophil ($1.8\text{--}6.3\times 10^9/\text{L}$) | 10.77 | 14.65 | 11.04 | 84.3 |
| Lymphocyte ($1.1\text{--}3.2\times 10^9/\text{L}$) | 0.55 ↓ | 0.75 ↓ | 0.53 ↓ | 1.21 |
| Eosinophils ($0.02\text{--}0.52\times 10^9/\text{L}$) | 0 ↓ | 0 ↓ | 0 ↓ | 0.02 |
| Red blood cell ($4.3\text{--}5.8\times 10^9/\text{L}$) | 4.18 ↓ | 3.67 ↓ | 3.93 ↓ | 3.63 ↓ |
| Hemoglobin (130–175 g/L) | 139 | 120 ↓ | 127 ↓ | 117 ↓ |
| C-reactive protein (0–5 mg/L) | 277.76 ↑ | 162.4 ↑ | 19.94 ↑ | 7.16 ↑ |
| Serum amyloid protein (0.1–10 mg/L) | 1843.8 ↑ | 980.75 ↑ | 232.65 ↑ | 99.09 ↑ |
| Procalcitonin (0.04–0.25 ng/mL) | 4.33 ↑ | 0.52 ↑ | 0.08 | - |
| Alanine aminotransferase (5–40 U/L ) | 56.4 ↑ | 59.8 ↑ | 98.9 ↑ | 64.8 ↑ |
| Aspartate aminotransferase (8–40 U/L) | 50 ↑ | 56.6 ↑ | 93.6 ↑ | 48 ↑ |
| Serum total protein (60–83 g/L ) | 67.7 | 49.1 ↓ | 55 ↓ | 55.5 ↓ |
| Serum albumin (34–54 g/L ) | 34.7 | 26.1 ↓ | 25.1 ↓ | 25.9 ↓ |
| Creatine kinase (25–200 U/L ) | 102 | 853 ↑ | 2476 ↑ | 441 ↑ |
| Lactic dehydrogenase (109–245 U/L) | 183 | 160 | 241 | 220 |
| Blood urea nitrogen (3–9.2 mmol/L) | 7.38 | 7.1 | 8.3 | 7.3 |
| Creatinine (44–120 $\mu\text{mol/L}$) | 78 | 86 | 78 | 68 |
| Serum iron (10.6–36.7 $\mu\text{mol/L}$) | 1.80 ↓ | 6.16 ↓ | 17.56 | 13.35 |

**Table 2.** **2019-nCoV RNA nasopharyngeal swab tests in the patient with COVID-19**

| 2019-nCoV<br>detection date | February 13 | February<br>19 | February<br>21 | February<br>23 | February<br>26 | February<br>28 |
| --- | --- | --- | --- | --- | --- | --- |
| Results | Positive | Negative | Negative | Positive | Negative | Negative |

**Figure 2.**
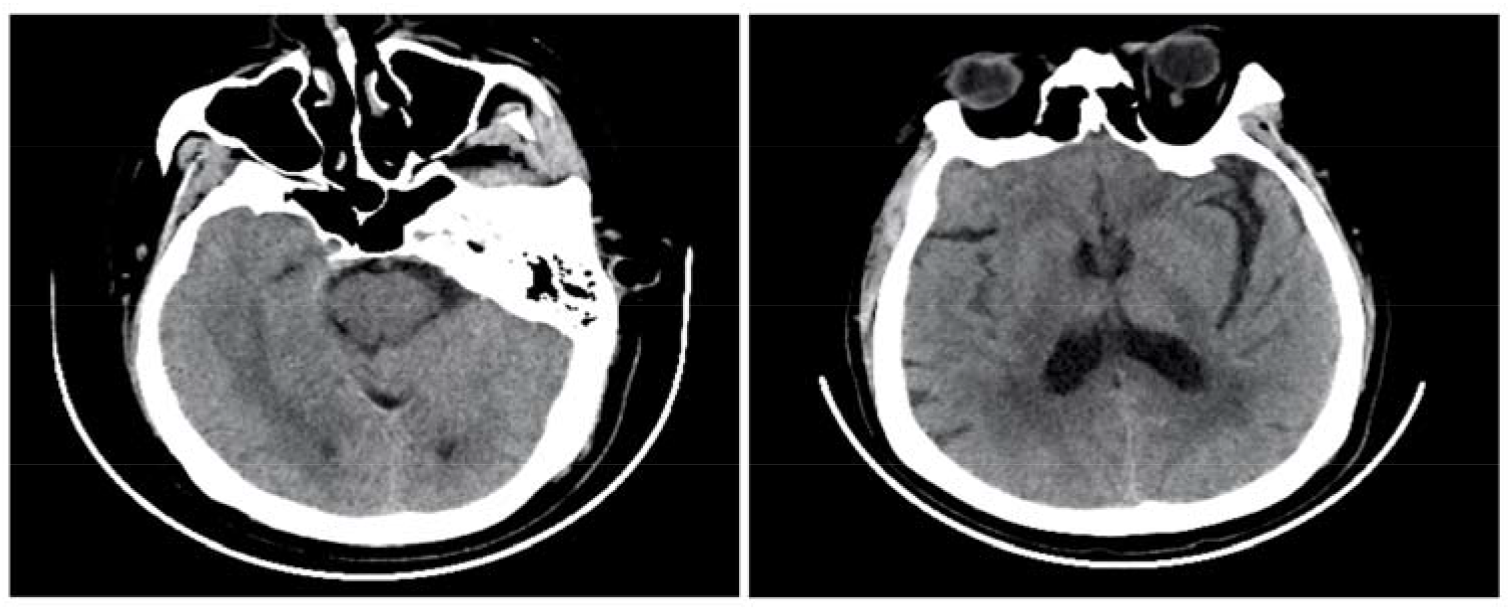
Cranial CT imaging of a patient with COVID-19. Cranial CT revealed bilateral basal ganglia and paraventricular lacunar infarction and brain atrophy. COVID-19, coronavirus disease 2019; CT, computed tomography

Based on the positive 2019-nCoV test result, he was diagnosed with COVID-19. Based on the acute flaccid myelitis of the lower limbs, urinary and bowel incontinence, and sensory level at T10, the diagnosis of acute myelitis was more likely. After admission, oxygen inhalation treatment with high-flow nasal catheters was administered. Meanwhile, the patient was received comprehensive drug therapies: ganciclovir (0.5 g once daily) for 14 days, lopinavir/ritonavir (500 mg twice daily) for 5 days, moxifloxacin (400 mg once daily) for 6 days, meropenem (1 g twice daily) for 8 days, glutathione (1.8 g once daily) for 12 days, dexamethasone (10 mg once daily) for 10 days, human immunoglobulin (15 g once daily) for 7 days, mecobalamin (1000 µg once daily) for 14 days, and pantoprazole (80 mg once daily) for 10 days. On the second day after admission, the patient’s body temperature returned to normal, and the oxygen saturation was greater than 93% at rest. There was no occurrence of adverse drug reactions or contraindications. His muscle strength in both upper limbs recovered to grade 4/5 and that in both lower limbs was grade 1/5. Two nCoV RNA nasopharyngeal swab test results were negative with an interval of more than one day. Re-examination of pulmonary CT showed that the lesions were absorbed and met the discharge criteria of COVID-19. He was then discharged and transferred to a designated hospital for isolation and rehabilitation therapy.

## Discussion

Our report is the first to present a case of post-infectious myelitis in the world, indicating that acute myelitis may be a neurological complication of COVID-19. Microbes including *M. pneumoniae*, EBV, CMV, rhinovirus, and measles are implicated in post-infectious acute myelitis^[11-13]^. *M. pneumoniae* may be one of the most widely recognized infections. A probable hypothesis is that infectious organisms are targeted by the immune system, which also attacks CNS tissue because of structural similarities between the microbial cellular wall components and neuronal receptors^[14]^. In this study, the IgM antibodies for common infectious organisms including *M. pneumoniae*, EBV, and CMV tested negative, and the symptoms of acute myelitis occurred after high fever and the diagnosis of COVID-19, suggesting that 2019-nCov might be the pathogenic virus. Moreover, a recent study showed that SARS-CoV-2 can enter the human body via ACE2 receptors on the surface of human cells and cause disease^[5]^. It is intriguing that ACE2 receptors are also expressed on the membrane of spinal cord neurons^[9,10]^, further suggesting that SARS-CoV-2 is implicated in acute myelitis based on the specific ACE2 receptors on the surface of spinal cord neurons.

Based on our study, acute paralysis is a novel neurological symptom of COVID-19. An analysis of the clinical characteristics of 1099 patients with COVID-19 showed that common symptoms at the onset of the disease are fever (on admission in 43.8% of patients, and after admission in 88.7% of patients) and cough (67.8%), while neurological symptoms are rare^[2]^. However, patients with severe COVID-19 are likely to develop neurological symptoms (such as headache, dizziness, hypogeusia, and neuralgia) and complications, including acute cerebrovascular diseases, impaired consciousness, and skeletal muscular injury^[15]^.

According to the laboratory testing, we found that lymphopenia and eosinopenia occurred in the early stage of COVID-19. The decrease in red blood cells and hemoglobin levels showed that the bone marrow hematopoietic system was affected by SARS-CoV-2, which is expected to be further confirmed by large-scale retrospective analysis. ACE2 is reportedly expressed in hepatic bile duct cells and proximal renal tubules^[7,8]^, and hepatic and renal dysfunctions are also detected in patients with COVID-19^[16]^. The liver function in our patient was obviously damaged with normal renal function, and elevated levels of alanine aminotransferase and aspartate aminotransferase reached a peak on the 15th day after disease onset, and then recovered gradually. Meanwhile, the levels of total protein, albumin, and HDL-C significantly decreased with disease progression, suggesting that metabolic abnormalities contributed to the pathogenesis of COVID-19. However, the renal function was normal in this study, which was inconsistent with that in a previous study. It has been reported that a decreased level of serum iron in patients with pneumonia is an independent risk factor for in-hospital death^[17]^. The mechanism may be related to the influence of pathogenic microorganism infection on iron uptake. In our patient, on the 7th day of disease onset, the serum iron level was 1.8 μmol/L and gradually increased with the treatment. The serum iron level returned to normal when the nucleic acid test result of 2019-nCov was negative. The possible role of serum iron in the 2019-nCov infection warrants further research in the future. We believe that an overactive inflammatory response and immune damage occurred in COVID-19 for high levels of C-reactive protein, serum amyloid A, interleukin-6, and ferritin in our patient. Immune damage and cytokines released by inflammatory storms in the early stage of COVID-19 might explain why the spinal cord was implicated in the disease.

## Conclusion

Our report is the first to present a COVID-19 case with spinal cord involvement 1 week after the onset of fever, wherein the clinical findings indicated post-infectious acute myelitis. Acute myelitis may be a neurological complication of COVID-19.

## Data Availability

No datasets were generated or analyzed for this study.

## Contributions

KZ and S-KN conceptualized the paper. S-KN analyzed the data with input from KZ, J-CH, and Y-WF. S-KN and L-ML wrote the initial draft, with all authors providing critical feedback and edits for subsequent revisions. All authors approved the final draft of the manuscript.

## Ethical approval

This study was approved by the Third People’s Hospital of Hubei Province.

## Patient consent

Consent was obtained from the patient.

